# Colonoscopy and reduction of colorectal cancer risk by molecular tumor subtypes: a population-based case-control study

**DOI:** 10.1101/2020.01.10.20017137

**Authors:** Michael Hoffmeister, Hendrik Bläker, Lina Jansen, Elizabeth Alwers, Efrat L. Amitay, Prudence R. Carr, Matthias Kloor, Esther Herpel, Wilfried Roth, Jenny Chang-Claude, Hermann Brenner

## Abstract

**Objective:** In previous studies, the protective effect of colonoscopy was generally stronger for distal than for proximal colorectal cancer (CRC). This study aimed to investigate whether the association of colonoscopy and CRC risk varies according to major molecular pathological features and pathways of CRC.

**Design:** Population-based case-control study from Germany, including 2132 patients with a first diagnosis of CRC and information on major molecular tumor markers, and 2486 control participants without CRC. Detailed participant characteristics were collected by standardized questionnaires. Information on previous colonoscopy was derived from medical records. Polytomous logistic regression was used to calculate adjusted odds ratios (ORs) and 95% confidence intervals (95% CI) for the association between previous colonoscopy and subtypes of CRC.

**Results:** Overall, we observed strong risk reduction of CRC after colonoscopy that was weaker for microsatellite instable (MSI) than for non-MSI CRC (p for heterogeneity <0.01), for CpG island methylator phenotype (CIMP) high CRC than for CIMP low/negative CRC (p het<0.01), for BRAF-mutated than for BRAF non-mutated CRC (p het<0.01), for KRAS non-mutated than for KRAS-mutated CRC (p het=0.04), and for CRC classified into the sessile serrated pathway than for CRC of the traditional pathway (p het<0.01). After colonoscopy with detection of adenomas, no risk reduction was found for sessile serrated pathway CRC, MSI and BRAF mutated subtypes.

**Conclusion:** Our study extends the molecular understanding of existing differences in risk reduction of proximal and distal CRC reported by previous studies, and may imply important information for improving strategies for timely detection of relevant precursors.

**Summary Box:** *What is already known about this subject?:*

- Colonoscopy is an effective tool not only for early detection but also for prevention of colorectal cancer.
- In previous studies, risk reduction after colonoscopy was generally stronger for cancer of the distal colon and rectum than for cancer of the proximal colon.

*What are the new findings?:*

- This observational study found variation of colorectal cancer risk reduction after colonoscopy according to major molecular subtypes characteristic of the proximal colon (MSI, CIMP-high, BRAF mutation), and for colorectal cancer potentially developing via the sessile serrated pathway.

*How might it impact on clinical practice in the foreseeable future?:*

- This study contributes to the identification of molecular characteristics and associated phenotypes of potentially missed or more aggressive precursors.
- The study provides important information for improving strategies for a timely detection of relevant precursors at colonoscopy.

## Introduction

In colorectal cancer (CRC), colonoscopy is an effective tool not only for early detection but also for prevention, because benign neoplastic precursor lesions, such as conventional adenomas and serrated polyps, can be detected and removed during the examination ^1-3^. In previous studies, the association of previous colonoscopies and risk reduction of CRC was investigated for CRC by subsite. In these studies, risk reduction was generally stronger for cancer of the distal colon and rectum than for cancer of the proximal colon ^2,4^.

CRC is a heterogeneous disease that develops via different pathways, and the different molecular pathological pathways are related to the detectability of precursors at colonoscopy and perhaps also to the rapidness of progression to cancer ^5^. For example, endoscopic detection of sessile serrated adenomas (also referred to as sessile serrated polyps or lesions) was shown to be highly variable in recent studies ^6,7^.

Traditional adenomas, sessile serrated adenomas and traditional serrated adenomas are distinct precursors with malignant potential which can develop into CRC. The underlying pathways can be characterized by molecular tumor markers including microsatellite instability (MSI), CpG island methylator phenotype (CIMP), and KRAS and BRAF mutations ^8^. Although prevalence of the molecular subtypes is often closely linked to anatomic subsites, molecular tumor characterization provides more specific information about underlying associations and pathways ^9^.

So far, studies have investigated previous colonoscopy and prevalence of single molecular subtypes among CRC cases ^10^, but no study has yet comprehensively investigated the potential of colonoscopy for risk reduction of CRC according to multiple major subtypes and pathways. Thus, we aimed to investigate whether the association of previous colonoscopy and CRC risk varies according to major molecular pathological markers and pathways of CRC.

## Methods

### Study design and study population

Patients with CRC and controls included in this study are participants of the DACHS study, a population-based case-control study in the southwest of Germany that has been ongoing since 2003. All 22 hospitals offering CRC surgery in the study region take part in the recruitment of patients. Patients and controls are eligible for this study if they are at least 30 years old (no upper age limit), are able to speak German and are also able to participate in an interview of about one hour. Only patients with a first diagnosis of primary CRC are eligible for this study. It has been estimated that approximately 50% of all eligible CRC patients in the study region of approximately two million inhabitants are recruited in this population-based study. Control participants are randomly drawn from population registries and frequency-matched to the patients by age, sex, and county of residence. Those with a previous diagnosis of CRC are excluded. Participation rate among community-based controls is 51% overall. Participation rates among cases and controls under age 75 are higher than in the oldest age group. More information on the study design has been reported previously ^3,11^.

In this analysis, only patients with information from molecular tumor analyses were included, which applied for patients recruited between 2003 and 2010. Control participants included in this analysis were recruited in the same time period. The study was approved by the ethical committees of the Medical Faculty of the University of Heidelberg and the Medical Chambers of Baden-Württemberg and Rhineland-Palatinate. Written informed consent was obtained from each participant.

### Data collection

The patients and controls provided information during a face-to-face interview. Control participants who declined participation in the face-to-face interview were offered a self-administered short questionnaire to assess key information. In addition, we collected discharge letters and tumor pathology reports, and reports of previous lower endoscopies of the patients and controls.

### Assessment of previous endoscopies

Information about previous lower endoscopies (not including endoscopies related to the diagnosis of CRC) was obtained in detail during the interviews. If previous lower endoscopies were reported, we requested the matching endoscopy reports from the participants’ physicians. In this analysis, participants were included for whom medical records of the last previous endoscopy corresponding to the last self-reported previous endoscopy could be obtained, regardless of the indication for colonoscopy. The year, type and result of endoscopy were derived from medical records. In addition, screening colonoscopies which detected CRC were also counted as previous lower endoscopy.

### Tumor tissue analyses

For 2,463 unselected patients, results from molecular tumor tissue analyses (MSI, BRAF, KRAS, CIMP, MLH1 methylation) were available. Details on collection, processing and marker analyses have been reported previously ^11-15^ and are provided in the **Supplementary Material**.

### Study exclusions

Among 2463 patients and 3145 controls, participants with inflammatory bowel disease (N=39) or with missing information on previous endoscopies were excluded (N=2) (**Supplementary Figure 1**). Also, participants were excluded if the endoscopy report of the last self-reported endoscopy was not obtained (N=604) or if the last endoscopy was not a colonoscopy (N=62). We further excluded participants if the last colonoscopy was incomplete (did not reach coecum) or if bowel preparation quality was impaired (N=239) according to the endoscopy report or if no information could be derived regarding adenoma removal (N=34).

### Statistical analyses

Characteristics of the study population were reported by case-control status (Table 1). CRCs developing via the different pathways were classified as suggested by Leggett et al. ^8^: traditional pathway (MSS, CIMP low/neg, BRAF non-mutated, KRAS non-mutated); sessile serrated adenoma pathway (BRAF mutated, CIMP-high); alternate pathway (MSS, KRAS mutated, non-CIMP high). The ‘all other pathways’ includes patients with full information on the four markers who could not be classified into either of the aforementioned pathways and were not included in pathway analyses (**Supplementary Table 1**).

**Table 1.** Characteristics of the study population.

|  | Cases (%) | Controls (%) | p-value |
| --- | --- | --- | --- |
| **Age** |  |  |  |
| <65 y | 692 (32%) | 769 (31%) | **0.0711** |
| 65-74 y | 756 (36%) | 963 (39%) |  |
| ≥75 y | 684 (32%) | 754 (30%) |  |
| **Female** | 891 (42%) | 1004 (40%) | 0.3330 |
| **School education** |  |  |  |
| <10 y | 1449 (68%) | 1483 (60%) | **<0.0001** |
| 10-11 y | 356 (17%) | 485 (20%) |  |
| ≥12 y | 323 (15%) | 498 (20%) |  |
| **1st degree family history of CRC** | 317 (15%) | 274 (11%) | **0.0001** |
| **Body mass index** |  |  |  |
| <25 kg/m<sup>2</sup> | 828 (39%) | 834 (34%) | **<0.0001** |
| 25-29 kg/m<sup>2</sup> | 885 (42%) | 1197 (48%) |  |
| ≥30 kg/m<sup>2</sup> | 410 (19%) | 444 (18%) |  |
| **Smoking** |  |  |  |
| Never | 993 (47%) | 1309 (53%) | **<0.0001** |
| Former | 801 (38%) | 886 (36%) |  |
| Current | 332 (15%) | 283 (11%) |  |
| **Regular use of NSAIDs** | 498 (24%) | 735 (30%) | **<0.0001** |
| **Ever use of HRT** | 251 (12%) | 431 (17%) | **<0.0001** |
| **Diabetes** | 385 (18%) | 347 (14%) | **0.0003** |
| **Health check-up participation** | 1769 (84%) | 2108 (88%) | **0.0001** |
| **Previous colonoscopy\*** | 406 (19%) | 1054 (42%) | **<0.0001** |

CRC=colorectal cancer, NSAIDs=non-steroidal anti-inflammatory drugs (including aspirin), HRT=hormone replacement therapy among women. Covariates with missing values were school education (24 participants), 1st degree family history of CRC (14) body mass index (20), smoking (14), regular use of NSAIDs (87), ever use of HRT (14), diabetes (59), and health check-up participation (94).
\*All self-reported previous colonoscopies were confirmed by medical records. Screening colonoscopy detected CRCs were counted as previous colonoscopy.

Polytomous logistic regression analyses were performed to calculate adjusted odds ratios (ORs) and 95% confidence intervals (CIs) for the association of previous colonoscopy and CRC risk by subtype (outcome levels defined as e.g. controls=0, MSS=1, MSI-high=2; controls=0, traditional pathway=1, sessile serrated pathway=2, alternate pathway=3). The models were adjusted for age, sex, school education (≤9, 10-11, >11 years), history of CRC in a first degree relative, body mass index (BMI), smoking (current, former, never), ever regular use of non-steroidal anti-inflammatory drugs (NSAIDs, 2+ times per week), ever use of hormone replacement therapy, history of diabetes, and participation in previous health check-ups ^3,16^. In addition to analyses of CRC at any subsite, we assessed subtype-specific associations for proximal and distal location (from splenic flexure) including rectum separately. Also, we assessed subtype-specific risk of CRC after previous negative colonoscopy (no conventional or sessile serrated adenomas detected and removed) and previous colonoscopy with detection and removal of adenomas (including sessile serrated adenomas) at last colonoscopy with and without consideration of the time interval since last colonoscopy (≤3/>3 years, ≤5/>5 years). Heterogeneity of associations between previous colonoscopy and CRC subtypes was tested for statistical significance in case-case analyses.

All analyses were performed with SAS, version 9.4 (SAS Institute Inc., Cary, NC). Statistical tests were two-sided using an alpha level of 0.05. The tables including forest plots were generated using the ‘forestplot’ package in RStudio, R version 3.5.2.

## Results

Overall, 2132 CRC patients with available information on the molecular tumor features and 2486 population-based control participants with no CRC were included in this study. Both patients and controls were 69 years on average and slightly more than 40% were female (**Table 1**). A previous colonoscopy was reported and confirmed for 19% of the patients and for 42% of the controls. Among the patients, UICC (Union for International Cancer Control) stages I, II, III, and IV were diagnosed in 18%, 35%, 33%, and 14%, respectively. Proximal cancers were more frequent among women (40%) than among men (30%).

In multivariable analyses, previous colonoscopy was associated with strong risk reduction of CRC (OR 0.32, 95% CI 0.27-0.36). Risk reduction was, however, weaker for MSI-high CRC (p for subtype heterogeneity (het) <0.0001), CIMP-high CRC (p=0.0041), BRAF mutated CRC (p het <0.0001), and sessile serrated pathway CRC (p het=0.0090), respectively. Risk reduction of KRAS mutated and non-mutated CRC differed only slightly (p het=0.0432) (**Table 2**). Cancers characterized as sessile serrated CRCs were by definition CIMP-high and BRAF mutated, but mostly showed MSI (74%) and MLH1 hypermethylation (72%), as well (**Supplementary Table 1**).

**Table 2.**
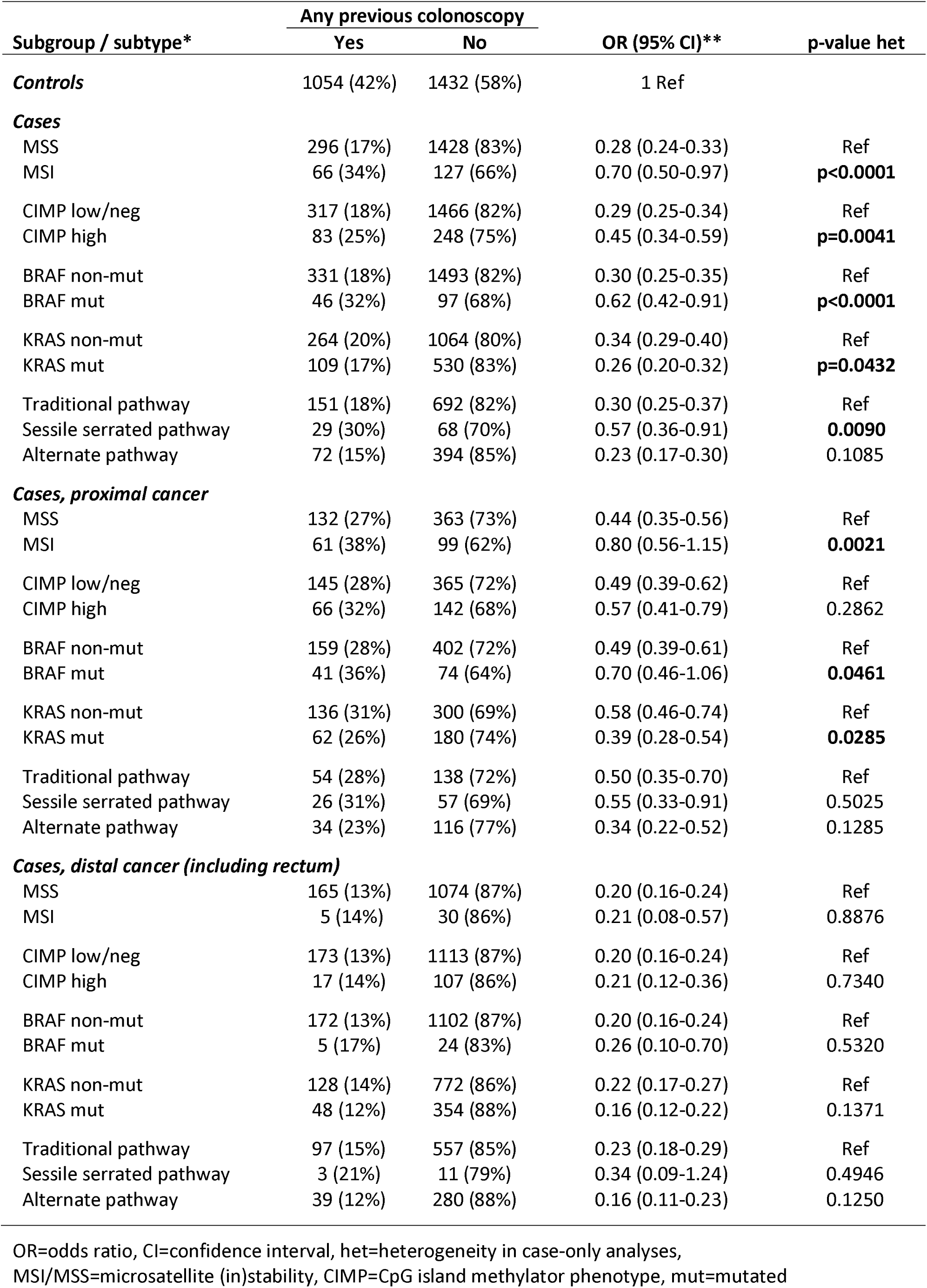
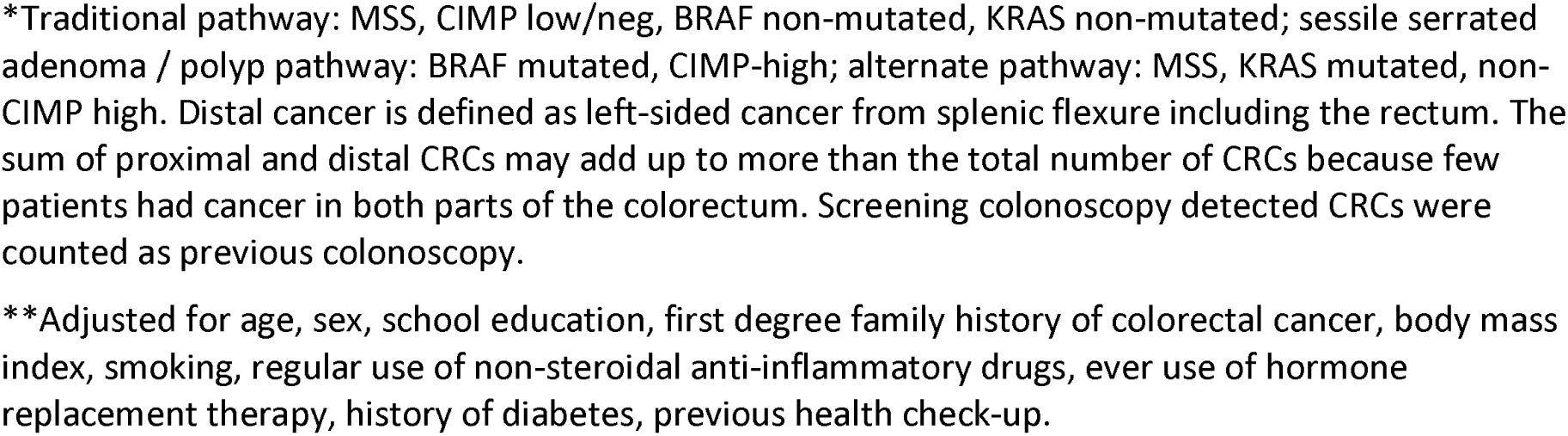
Risk of colorectal cancer subtypes after previous colonoscopy at any site, and by location of colorectal cancer.

| Subgroup / subtype* | Any previous colonoscopy |  | OR (95% CI)** | p-value het |
| --- | --- | --- | --- | --- |
|  | Yes | No |  |  |
| **Controls** | 1054 (42%) | 1432 (58%) | 1 Ref |  |
| **Cases** |  |  |  |  |
| MSS | 296 (17%) | 1428 (83%) | 0.28 (0.24-0.33) | Ref |
| MSI | 66 (34%) | 127 (66%) | 0.70 (0.50-0.97) | **p<0.0001** |
| CIMP low/neg | 317 (18%) | 1466 (82%) | 0.29 (0.25-0.34) | Ref |
| CIMP high | 83 (25%) | 248 (75%) | 0.45 (0.34-0.59) | **p=0.0041** |
| BRAF non-mut | 331 (18%) | 1493 (82%) | 0.30 (0.25-0.35) | Ref |
| BRAF mut | 46 (32%) | 97 (68%) | 0.62 (0.42-0.91) | **p<0.0001** |
| KRAS non-mut | 264 (20%) | 1064 (80%) | 0.34 (0.29-0.40) | Ref |
| KRAS mut | 109 (17%) | 530 (83%) | 0.26 (0.20-0.32) | **p=0.0432** |
| Traditional pathway | 151 (18%) | 692 (82%) | 0.30 (0.25-0.37) | Ref |
| Sessile serrated pathway | 29 (30%) | 68 (70%) | 0.57 (0.36-0.91) | **0.0090** |
| Alternate pathway | 72 (15%) | 394 (85%) | 0.23 (0.17-0.30) | 0.1085 |
| **Cases, proximal cancer** |  |  |  |  |
| MSS | 132 (27%) | 363 (73%) | 0.44 (0.35-0.56) | Ref |
| MSI | 61 (38%) | 99 (62%) | 0.80 (0.56-1.15) | **0.0021** |
| CIMP low/neg | 145 (28%) | 365 (72%) | 0.49 (0.39-0.62) | Ref |
| CIMP high | 66 (32%) | 142 (68%) | 0.57 (0.41-0.79) | 0.2862 |
| BRAF non-mut | 159 (28%) | 402 (72%) | 0.49 (0.39-0.61) | Ref |
| BRAF mut | 41 (36%) | 74 (64%) | 0.70 (0.46-1.06) | **0.0461** |
| KRAS non-mut | 136 (31%) | 300 (69%) | 0.58 (0.46-0.74) | Ref |
| KRAS mut | 62 (26%) | 180 (74%) | 0.39 (0.28-0.54) | **0.0285** |
| Traditional pathway | 54 (28%) | 138 (72%) | 0.50 (0.35-0.70) | Ref |
| Sessile serrated pathway | 26 (31%) | 57 (69%) | 0.55 (0.33-0.91) | 0.5025 |
| Alternate pathway | 34 (23%) | 116 (77%) | 0.34 (0.22-0.52) | 0.1285 |
| **Cases, distal cancer (including rectum)** |  |  |  |  |
| MSS | 165 (13%) | 1074 (87%) | 0.20 (0.16-0.24) | Ref |
| MSI | 5 (14%) | 30 (86%) | 0.21 (0.08-0.57) | 0.8876 |
| CIMP low/neg | 173 (13%) | 1113 (87%) | 0.20 (0.16-0.24) | Ref |
| CIMP high | 17 (14%) | 107 (86%) | 0.21 (0.12-0.36) | 0.7340 |
| BRAF non-mut | 172 (13%) | 1102 (87%) | 0.20 (0.16-0.24) | Ref |
| BRAF mut | 5 (17%) | 24 (83%) | 0.26 (0.10-0.70) | 0.5320 |
| KRAS non-mut | 128 (14%) | 772 (86%) | 0.22 (0.17-0.27) | Ref |
| KRAS mut | 48 (12%) | 354 (88%) | 0.16 (0.12-0.22) | 0.1371 |
| Traditional pathway | 97 (15%) | 557 (85%) | 0.23 (0.18-0.29) | Ref |
| Sessile serrated pathway | 3 (21%) | 11 (79%) | 0.34 (0.09-1.24) | 0.4946 |
| Alternate pathway | 39 (12%) | 280 (88%) | 0.16 (0.11-0.23) | 0.1250 |

OR=odds ratio, CI=confidence interval, het=heterogeneity in case-only analyses, MSI/MSS=microsatellite (in)stability, CIMP=CpG island methylator phenotype, mut=mutated
\*Traditional pathway: MSS, CIMP low/neg, BRAF non-mutated, KRAS non-mutated; sessile serrated adenoma / polyp pathway: BRAF mutated, CIMP-high; alternate pathway: MSS, KRAS mutated, non-CIMP high. Distal cancer is defined as left-sided cancer from splenic flexure including the rectum. The sum of proximal and distal CRCs may add up to more than the total number of CRCs because few patients had cancer in both parts of the colorectum. Screening colonoscopy detected CRCs were counted as previous colonoscopy.
\*\*Adjusted for age, sex, school education, first degree family history of colorectal cancer, body mass index, smoking, regular use of non-steroidal anti-inflammatory drugs, ever use of hormone replacement therapy, history of diabetes, previous health check-up.

In the proximal colon, risk reduction of CRC was overall weaker than in the distal colorectum. Also, differences in risk reduction according to subtypes were more pronounced for proximal than for distal CRC. In the proximal colon, we observed weaker or no risk reduction for MSI (OR 0.80, 0.56-1.15; MSS: OR 0.44, 0.35-0.56; p het=0.0021), BRAF-mutated (OR 0.70, 0.46-1.06; BRAF non-mutated: OR 0.49, 0.39-0.61; p het=0.0461), and KRAS non-mutated CRC (OR 0.58, 0.46-0.74; KRAS-mutated: OR 0.39, 0.28-0.54; p het=0.0285).

MSI-high CRC, BRAF-mutated CRC, and sessile serrated pathway CRC were less frequent in the distal colorectum. Differences in risk reduction after colonoscopy seemed to be minor for the investigated tumor characteristics in the distal colorectum.

In analyses on previous colonoscopy with no detection of adenomas versus no previous colonoscopy, colonoscopy itself is not actively reducing the risk of CRC (a group with low risk of CRC is compared to a group at average risk of CRC). Risk reduction was less pronounced for MSI-high CRC than for MSS CRC (p het=0.0001) and for BRAF-mutated compared with BRAF non-mutated CRC (p het <0.0001) (**Table 3**). When differentiating by time since last colonoscopy without detection of adenomas (**Figure 1A**), risk reduction was less pronounced for BRAF-mutated CRC if colonoscopy was conducted ≤3 years before, and for MSI CRC, CIMP-high CRC, BRAF mutated CRC and sessile serrated pathway CRC if the colonoscopy was more than 3 years ago.

**Table 3.** Risk of colorectal cancer subtypes after previous_colonoscopy <u>with no</u> detection of adenomas.

| Subgroup / subtype* | Last colonoscopy negative | No previous colonoscopy | OR (95% CI)** | p-value het |
| --- | --- | --- | --- | --- |
| **Controls** | 830 (37%) | 1432 (63%) | 1 Ref |  |
| **Cases** |  |  |  |  |
| MSS | 103 (7%) | 1428 (93%) | 0.12 (0.09-0.15) | Ref |
| MSI | 26 (17%) | 127 (83%) | 0.32 (0.20-0.50) | **0.0001** |
| CIMP low/neg | 120 (8%) | 1466 (92%) | 0.13 (0.10-0.16) | Ref |
| CIMP high | 27 (10%) | 248 (90%) | 0.17 (0.11-0.26) | 0.2283 |
| BRAF non-mut | 118 (7%) | 1493 (93%) | 0.12 (0.10-0.15) | Ref |
| BRAF mut | 23 (19%) | 97 (81%) | 0.36 (0.22-0.59) | **<0.001** |
| KRAS non-mut | 102 (9%) | 1064 (91%) | 0.15 (0.12-0.19) | Ref |
| KRAS mut | 37 (7%) | 530 (93%) | 0.11 (0.08-0.16) | 0.1325 |
| Traditional pathway | 48 (6%) | 692 (94%) | 0.11 (0.08-0.16) | Ref |
| Sessile serrated pathway | 10 (13%) | 68 (87%) | 0.23 (0.11-0.45) | 0.0899 |
| Alternate pathway | 25 (6%) | 394 (94%) | 0.10 (0.06-0.15) | 0.6423 |

OR=odds ratio, CI=confidence interval, het=heterogeneity in case-only analyses, MSI/MSS=microsatellite (in)stability, CIMP=CpG island methylator phenotype, mut=mutated
\*\*Adjusted for age, sex, school education, first degree family history of colorectal cancer, body mass index, smoking, regular use of non-steroidal anti-inflammatory drugs, ever use of hormone replacement therapy, history of diabetes, previous health check-up.

**Figure 1.**
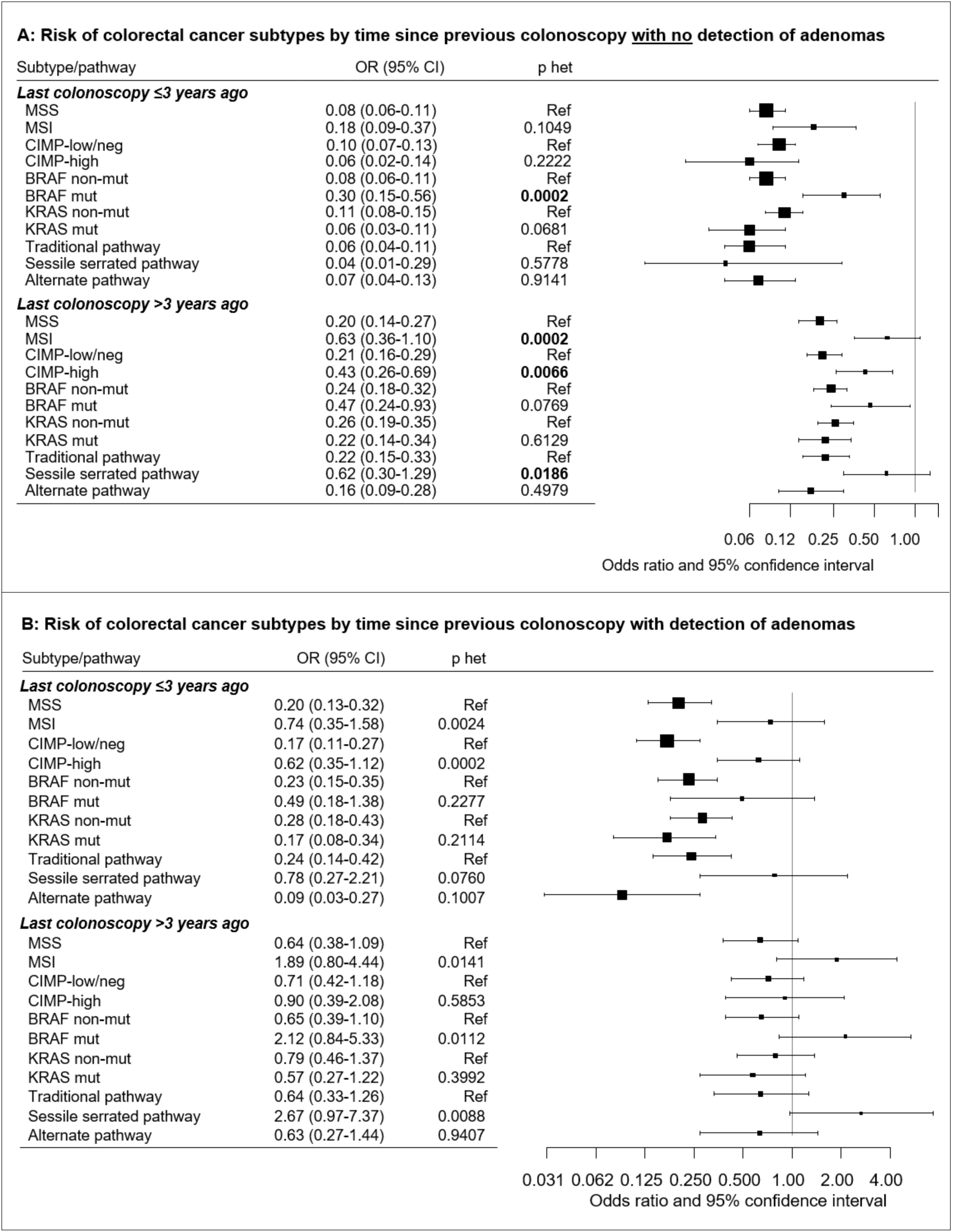
Risk of colorectal cancer subtypes according to time since last colonoscopy (≤3 years / >3 years) compared to no previous endoscopy. A: Last colonoscopy with no detection of adenomas; B: Last colonoscopy with detection of adenomas (OR=odds ratio, CI=confidence interval, MSI/MSS=microsatellite (in)stability, CIMP=CpG island methylator phenotype, mut=mutated. Traditional pathway: MSS, CIMP low/neg, BRAF non-mutated, KRAS non-mutated; sessile serrated adenoma / polyp pathway: BRAF mutated, CIMP-high; alternate pathway: MSS, KRAS mutated, non-CIMP high). Scales on the x-axis had to be different in A and B for better readability.

In analyses on previous colonoscopy with detection of adenomas, risk reduction of CRC was overall weaker than after colonoscopy with no detection of adenomas as expected (**Table 4**). However, much lower or no risk reduction or no risk reduction was observed for MSI CRC (OR 1.02, 0.57-1.81, p het=0.0002), CIMP-high CRC (OR 0.69, 0.42-1.12, p het=0.0024), BRAF mutated CRC (OR 0.90, 0.45-1.80, p het=0.0086), and CRC with sessile serrated pathway features (OR 1.24, 0.59-2.62, p het=0.0025). When the last colonoscopy with detection of adenomas was ≤3 years before, risk reduction of CRC was weaker or not observed if tumors were MSI, CIMP-high, BRAF-mutated, or classified into the sessile serrated pathway (**Figure 1B**). If the last colonoscopy with detection of any adenoma was more than 3 years ago, risk of CRC was up to more than twofold increased for MSI, BRAF mutated, and sessile serrated carcinomas and only moderately or not reduced for other subtypes. Additional results differentiating between colonoscopies conducted ≤5 and >5 years before are reported in **Supplementary Figure 2**.

**Table 4.** Risk of colorectal cancer subtypes after previous colonoscopy <u>with</u> detection of adenomas.

| Subgroup / subtype* | Adenoma(s)<br>at last<br>colonoscopy | No<br>colonoscopy | OR (95% CI)** | p-value het |
| --- | --- | --- | --- | --- |
| **Controls** | 159 (10%) | 1432 (90%) | 1 Ref |  |
| **Cases** |  |  |  |  |
| MSS | 53 (4%) | 1428 (96%) | 0.31 (0.22-0.43) | Ref |
| MSI | 16 (12%) | 127 (88%) | 1.02 (0.57-1.81) | **0.0002** |
| CIMP low/neg | 53 (3%) | 1466 (97%) | 0.30 (0.21-0.42) | Ref |
| CIMP high | 21 (8%) | 248 (92%) | 0.69 (0.42-1.12) | **0.0024** |
| BRAF non-mut | 61 (4%) | 1493 (96%) | 0.33 (0.24-0.46) | Ref |
| BRAF mut | 10 (9%) | 97 (91%) | 0.90 (0.45-1.80) | **0.0086** |
| KRAS non-mut | 49 (4%) | 1064 (96%) | 0.40 (0.28-0.56) | Ref |
| KRAS mut | 20 (4%) | 530 (96%) | 0.26 (0.16-0.44) | 0.1398 |
| Traditional pathway | 27 (4%) | 692 (96%) | 0.34 (0.22-0.51) | Ref |
| Sessile serrated pathway | 9 (12%) | 68 (88%) | 1.24 (0.59-2.62) | **0.0025** |
| Alternate pathway | 12 (3%) | 394 (97%) | 0.22 (0.11-0.41) | 0.2217 |

OR=odds ratio, CI=confidence interval, het=heterogeneity in case-only analyses, MSI/MSS=microsatellite (in)stability, CIMP=CpG island methylator phenotype, mut=mutated
\*Traditional pathway: MSS, CIMP low/neg, BRAF non-mutated, KRAS non-mutated; sessile serrated adenoma / polyp pathway: BRAF mutated, CIMP-high; alternate pathway: MSS, KRAS mutated, non-CIMP high.
\*\*Adjusted for age, sex, school education, first degree family history of colorectal cancer, body mass index, smoking, regular use of non-steroidal anti-inflammatory drugs, ever use of hormone replacement therapy, history of diabetes, previous health check-up.

In sensitivity analyses with restriction of participants with a previous colonoscopy to those with only one previous colonoscopy, odds ratios and tests for heterogeneity were very similar (data not shown).

## Discussion

In this study on the protective effect of a previous colonoscopy, we found variation in risk reduction of CRC according to molecular pathological subtypes. In particular, despite a generally strong reduction of CRC risk, risk reduction was less pronounced for tumors showing MSI, BRAF mutation or absence of KRAS mutation in the proximal colon. Only minor differences were observed in the distal colorectum. Also, risk reduction was weaker for CRC classified into the sessile serrated pathway (MSI/MSS, BRAF mutation, CIMP-high), especially after colonoscopy with detection of adenomas (including sessile serrated adenomas). This study provides a molecular basis for the currently discussed limitations of colonoscopy in reducing the risk of CRC in the proximal colon and for the molecular characterization of interval CRCs.

Our study is in line with previous studies on interval cancers regarding weaker risk reduction for MSI CRC, CIMP-high CRC and KRAS non-mutated CRC after colonoscopy^2,5,10,17,18^. A registry-based study among 566 CRC cases from Denmark found that post-colonoscopy CRCs were more often located in the proximal colon and were more often mismatch repair-deficient compared to those after no previous colonoscopy ^10^, and a more frequent post-colonoscopy occurrence of both MSI and CIMP-high CRCs was reported in a sub-analysis of two large prospective cohort studies from the United States ^2^. In four studies comparing interval with non-interval cancers, MSI and CIMP-high were more frequent in interval cancers, BRAF mutation frequency was not different, and KRAS mutation was less frequent in interval cancers compared with non-interval CRCs ^5,17-19^. Contrary to one study by Shaukat et al, colonoscopy associated risk reduction of BRAF mutated CRC was weaker in our study ^19^. In support of the latter finding, and in line with the analysis on KRAS by Shaukat ^18^, protection more pronounced for KRAS-mutated CRC in the proximal colon in our study. As BRAF-mutated and KRAS-mutated CRCs are mutually exclusive, the weaker protection against KRAS non-mutated CRC might partly be explained by prevalent BRAF-mutation. The molecular patterns reported for interval cancers and the patterns in risk reduction of single CRC subtypes observed in our study are characteristic of CRCs in the proximal colon.

For the first time, combined features based on the suggested pathways to CRC ^8^ were investigated. In most of our analyses, we observed less protection after previous colonoscopy for CRCs potentially arising via the sessile serrated pathway, and even no risk reduction was observed after a previous colonoscopy with adenoma detection. Sessile serrated adenomas represent about 15% of all removed polyps and are harder to detect at colonoscopy than traditional adenomas or traditional serrated adenomas, as sessile serrated adenomas are mostly flat or sessile, less bulky, and located in the proximal colon ^20-22^. Also, previous studies reported detection of sessile serrated adenomas to be highly variable between colonoscopy providers.^7^ Therefore, it is assumed that the higher proportion of sessile serrated CRC in interval cancers is due to missed precursors at colonoscopy ^23^. Another reason might be that sessile serrated adenomas are more aggressive and transform faster into CRC ^24^. Studies showed that size, dysplasia and inactivation by MLH1 seem to a play a role, and might be age-related, because MLH1 inactivation is more frequent in sporadic CRCs of older patients with MSI CRC ^22,23,25^.

Unfortunately, we had no information about the histological features of the polyps removed at previous colonoscopy. However, the lack of protection against CRC potentially arising from the sessile serrated pathway after previous colonoscopy with adenoma detection may point to the presence of multiple adenomas at previous colonoscopy, at which the less apparent sessile serrated adenomas may have been overlooked. The presence of multiple polyps has been reported as a risk factor for missed polyps at colonoscopy ^26,27^. Risk reduction of CRC potentially arising from the alternate pathway was mostly similar to risk reduction of CRC arising from the traditional pathway.

In previous analyses of this study, and in other observational studies, risk reduction through colonoscopy was stronger for distal than for proximal CRC ^2-4,16,28^. Other studies, however, even found no risk reduction after colonoscopy for either advanced neoplasms or CRC in the proximal colon ^29,30^. In the present study, we found that risk reduction after colonoscopy was not as strong for molecular subtypes characteristic of the proximal colon, i.e. MSI, CIMP-high, BRAF mutated CRC and CRCs potentially arising from the sessile serrated pathway compared to their paired subtypes. In particular, the weaker risk reduction observed for these subtypes was mostly due to weaker risk reduction in the proximal colon. This study included patients diagnosed between 2003 and 2010 and referred to colonoscopies conducted before diagnosis among CRC cases. At that time, guidelines started to classify sessile serrated polyps as CRC precursors, and many of them may not have been removed during colonoscopy before this was common practice, eventually contributing to a weaker risk reduction in the proximal colon. New strategies, more accurate classification and more awareness among gastroenterologists and pathologists are needed to improve detection of sessile serrated adenomas in the future ^31^. If molecular characteristics in CRC correspond to molecular characteristics of CRC precursors they may indicate more or less aggressive subtypes and might be used in addition to histological features to classify precursors. Thus, our data could be useful for the design of surveillance strategies as suggested by the analyses differentiating by time since last colonoscopy.

A number of strengths and limitations need to be considered in the interpretation of our population-based study. The analyses were performed in a large case-control study with comprehensive characterization of patients and control participants, and we were able to adjust for most known risk and protective factors of CRC. Detailed information about previous colonoscopy and potential adenoma detection were derived from medical records. Furthermore, the large number of patients for whom major molecular tumor markers were determined enabled us to perform statistically meaningful analyses on single tumor markers and pathway derived subtypes. The findings of this study are novel because no other study has yet comprehensively investigated risk reduction of CRC by multiple tumor markers and pathways. The main limitations of the study are related to its case-control design. In particular, selection bias in the recruitment of controls may have led to some overestimation of colonoscopy effects ^3^ which were somewhat stronger than those reported in other studies, especially cohort studies ^4^. Still, differences in colonoscopy-associated risk reduction and observed heterogeneity between subtypes of CRC are unlikely to be affected by such design issues, as they were derived from case-case analyses. The colonoscopies in this study were conducted before 2010, many even before the year 2000. Colonoscopy technique and quality have improved over time, and likewise the detection of precursors, which may have increased the protective effect of colonoscopies in more recent years. Finally, despite comprehensive adjustment for major potential confounding factors, residual confounding may still affect our results.

In conclusion, our study provides an empirical basis for the variation of risk reduction of CRC after colonoscopy according to major subtypes and pathways characteristic of the proximal colon (MSI, CIMP-high, BRAF mutation), and for sessile serrated CRC. This study extends the molecular understanding of differences in risk reduction of proximal and distal CRC consistently reported by previous studies, and describes molecular characteristics and associated phenotypes of potentially missed or more aggressive precursors for future improvement of colonoscopy.

## Data Availability

The data that support the findings of this study are available on request from the corresponding author.

## Acknowledgements

We are grateful to the study participants and the interviewers who collected the data. We would like to thank the following hospitals and cooperating institutions which recruited patients for this study: Chirurgische Universitätsklinik Heidelberg, Klinik am Gesundbrunnen Heilbronn, St Vincentiuskrankenhaus Speyer, St Josefskrankenhaus Heidelberg, Chirurgische Universitätsklinik Mannheim, Diakonissenkrankenhaus Speyer, Krankenhaus Salem Heidelberg, Kreiskrankenhaus Schwetzingen, St Marienkrankenhaus Ludwigshafen, Klinikum Ludwigshafen, Stadtklinik Frankenthal, Diakoniekrankenhaus Mannheim, Kreiskrankenhaus Sinsheim, Klinikum am Plattenwald Bad Friedrichshall, Kreiskrankenhaus Weinheim, Kreiskrankenhaus Eberbach, Kreiskrankenhaus Buchen, Kreiskrankenhaus Mosbach, Enddarmzentrum Mannheim, Kreiskrankenhaus Brackenheim, Cancer Registry of Rhineland-Palatinate, Mainz.

We are also very grateful for the support of the pathologies in the provision of tumor samples: Institut für Pathologie, Universitätsklinik Heidelberg; Institut für Pathologie, Klinikum Heilbronn; Institut für Angewandte Pathologie, Speyer; Pathologisches Institut, Universitätsklinikum Mannheim; Institut für Pathologie, Klinikum Ludwigshafen; Institut für Pathologie, Klinikum Stuttgart; Institut für Pathologie, Klinikum Ludwigsburg. Special thanks to the tissue bank of National Center for Tumor Diseases (NCT), Heidelberg, for storage and processing of the tissue samples.

We would like to thank Ute Handte-Daub, Ansgar Brandhorst, Dr Utz Benscheid, Bettina Walter, Gloria Laukemper, and Terence Osere for their excellent technical assistance.

## Abbreviations

CI: confidence interval
CIMP: CpG island methylator phenotype
CRC: colorectal cancer
het: heterogeneity
HRT: hormone replacement therapy
MSI: microsatellite instability
MSS: microsatellite stable
mut: mutated
NSAIDs: non-steroidal anti-inflammatory drugs
OR: odds ratio
UICC: Union for International Cancer Control

